# Clonal Hematopoiesis of Indeterminate Potential and the Risk of Cognitive Impairment in the Women’s Health Initiative Memory Study

**DOI:** 10.1101/2025.08.02.25332871

**Authors:** Yasminka A Jakubek, Aaron P Smith, Xiaoyan I Leng, Megan E Hall, Daniel Ezzat, Yash Pershad, Jason M Collins, Md Mesbah Uddin, David W Fardo, Pradeep Natarajan, Alexander G Bick, Jacob O Kitzman, Michael C Honigberg, Kathleen M Hayden, JoAnn E Manson, Siddhartha Jaiswal, Eric A Whitsel, Alexander P Reiner

## Abstract

**INTRODUCTION:** Clonal hematopoiesis of indeterminate potential (CHIP) confers an increased risk of several chronic aging-related diseases. Paradoxically, CHIP was associated with lower risk of dementia in recent studies.

**METHODS:** We examined associations between baseline CHIP and incident mild cognitive impairment (MCI) and/or probable dementia in the Women’s Health Initiative Memory Study. CHIP was detected using blood-based targeted sequencing. Cox proportional hazards models examined time to onset of cognitive impairment, adjusting for traditional risk factors.

**RESULTS:** Using a conventional variant allele fraction (VAF) threshold of 2%, CHIP was not associated with incident cognitive impairment. The presence of larger CHIP clone (VAF≥8%) was associated with a lower incidence of adjudicated probable dementia (HR=0.62 [95% CI=0.41-0.94], *p*=0.025), while the association with the composite outcome MCI/probable dementia was weaker and overlapped 1.0.

**DISCUSSION:** The association of CHIP with lower risk of cognitive impairment in postmenopausal women may be dependent on VAF and impairment severity.

## Introduction

Clonal hematopoiesis occurs when a hematopoietic cell acquires a somatic mutation and then undergoes clonal expansion, resulting in a genetically distinct leukocyte subpopulation. Clonal hematopoiesis of indeterminate potential (CHIP), a common age-related condition, is a type of clonal hematopoiesis defined by mutations in genes commonly mutated in myeloid leukemia[1]. Mutations in CHIP genes are associated with alterations in DNA methylation, DNA damage response, inflammation, malignant transformation, and other aging-related processes[2]. Emerging evidence suggests that CHIP is associated with acceleration of vascular disease processes including atherosclerosis, risk of cardiovascular events, and death[3–6].

Risk factors for Alzheimer’s disease (AD) and other forms of dementia include conditions associated with CHIP, such as inflammation, cardiovascular disease (CVD), metabolic disorders, and stroke[7]. Because CHIP appears to be an independent and potent CVD risk factor, it was originally hypothesized that CHIP would increase the risk for cognitive decline and dementia, either directly or indirectly, through cardiovascular pathways. Surprisingly, a recent set of detailed analyses of multiple cohorts suggested that CHIP is protective against AD, contrary to the original hypothesis[8]. A subsequent longitudinal analysis among patients with chronic kidney disease (CKD) similarly showed CHIP was associated with a lower risk of cognitive impairment[9].

The Women’s Health Initiative Memory Study (WHIMS) provides an opportunity to examine potential associations of CHIP with cognitive outcomes over as long as twenty-five years of observation. Herein we examine the association between CHIP and mild cognitive impairment (MCI) and probable dementia from WHIMS. We hypothesized that CHIP would be inversely associated with the risk of incident cognitive impairment. Secondary objectives were to explore the associations between specific driver mutations for CHIP and the incidence of cognitive impairment.

## Methods

### Participants and Setting

WHIMS was designed to evaluate the effects of conjugated equine estrogens alone (E-alone; in women with hysterectomy), or estrogen (E) in combination with medroxyprogesterone acetate (progestin; [E + P]), compared to placebo groups on the incidence of MCI or probable dementia[10]. All women provided written informed consent to participate. In total, 7,429 women from 39 sites of the Women’s Health Initiative (WHI) randomized trial for hormone therapy were enrolled in WHIMS between May 28, 1996, and December 13, 1999. All WHIMS participants were aged 65+ at baseline and met the inclusion criteria for the WHI hormone trials. The trials ended in 2002 (E + P) and 2004 (E-alone). In 2008, data collection transitioned from face-to-face clinic visits to telephone based cognitive assessments in the WHI Epidemiology of Cognitive Health Outcomes (WHIMS-ECHO)[11]. WHIMS-ECHO data collection continued to 2021 allowing for up to 26 years of observation (1995-2021). Cognitive status was adjudicated as MCI, probable dementia, or no impairment. Details of the cognitive assessment protocols and adjudication for WHIMS and WHIMS-ECHO are described in the **Supplement**.

In the current CHIP study, the cognitive impairment outcome was defined in two ways: (1) a combined outcome that includes incident MCI, adjudicated probable dementia, and self-reported dementia (i.e., combined MCI/dementia) during follow-up or (2) a more restrictive definition of dementia that includes only adjudicated probable dementia during follow-up (i.e., adjudicated probable dementia only), excluding individuals with MCI and those with dementia based only on self-report.

### CHIP assay and Definition of CHIP Status at Baseline

CHIP was assayed from blood in a subset of 4,949 WHIMS women as described in the **Supplement**. CHIP targeted sequencing and variant calling of the WHIMS samples were performed in two distinct batches with median read depths 5,791x (n=3,012) and 4,609x (n=1,955), respectively. Therefore, a covariate indicating sequencing batch was included in all CHIP association analyses.

In our primary analyses, CHIP was analyzed in two ways. (1) We defined CHIP as VAF ≥2% in one or more CHIP driver genes. (2) We also performed analyses in which CHIP carriers were limited to those with large CHIP clones (defined as VAF≥8%), to allow greater comparability to prior analyses of CHIP and AD conducted by Bouzid et al[8]. We also conducted sensitivity analyses modeling CHIP status as a categorical variable (<2%, 2-8%, >8%). In additional exploratory analyses, we defined CHIP status according to driver gene subtype (*DNMT3A, TET2*, other, or multiple), with each category being mutually exclusive.

### Covariates

Covariates included demographic and design factors at baseline: age, race and ethnicity, education, treatment assignment (E-alone, E+P, or placebo), and U.S. recruitment region (Midwest, Northeast, South, and West). Clinical covariates included body mass index (BMI; kg/m^2^), alcohol use (≥1 drinks/month), smoking history (never/former/current), self-reported history of diabetes, cholesterol-lowering medication, antihypertensive medication, and history of CVD (including myocardial infarction, stroke, heart failure, transient ischemic attack or TIA, coronary revascularization, peripheral artery disease, and carotid artery disease).

*APOE* genotypes were based on two SNPs, rs429358 and rs7412 that were available in a subsample of 4,508 participants. Genetic data were imputed and harmonized across WHI genome-wide association studies using the 1000 Genomes Project reference panel and MaCH algorithms implemented in Minimac[12]. Both SNPs had high imputation quality (R^2^ >0.97).

### Statistical Analysis

Baseline age was defined as age at blood draw. For both cognitive impairment outcomes, those who self-reported moderate or severe memory problems, dementia, AD, or blood cancer at or before the time of the blood draw used for CHIP analysis were excluded from downstream analyses (N = 8).

The association of baseline CHIP status (present versus absent) with cognitive impairment during follow-up was analyzed using Cox proportional hazards models with age as the underlying timescale. For the survival analyses, the onset of cognitive impairment was defined as time to MCI or time to dementia, as some participants did not have MCI prior to dementia. Each combination of CHIP VAF (exposure) and time to cognitive impairment (outcome) were tested with a set of three Cox regression models with increasing numbers of covariates. The first model (M1) included age at blood draw, sequencing batch, hormone therapy treatment assignment, race/ethnicity, and US region. The second model (M2) included all covariates from M1 as well as education level, baseline alcohol use, smoking history, BMI, cholesterol levels, hypertension, and CVD. CVD was a categorical variable with 3 groups: no CVD, stroke/TIA, or other CVD. The third model (M3) included all covariates from M1 and M2, as well as the presence of one or more APOE2 alleles, and the presence of one or more APOE4 alleles. For each model, individuals were included in the analysis if they had complete information for all covariates (n = 4934 for M1, n = 4655 for M2, and n = 4240 for M3). Baseline covariate comparisons between individuals with and without CHIP utilized Fisher’s exact test for categorical variables, t-tests with unequal variance for continuous variables, and normal approximated rate-ratio tests for crude incidence rates. Adjusted hazard ratios (HR) and 95% confidence intervals (CI) are reported for survival analyses. In additional sensitivity analysis, a Fine-Gray competing risk regression model was used to evaluate the association between CHIP and cognitive outcomes with death as the competing risk and adjusting for covariates as in the survival analysis. All statistical tests were two-sided with a significance threshold of 0.05 and computed in R version 4.2.0.

## Results

**Table 1** shows the demographic characteristics of the sample by CHIP status for 4,934 women (mean age 70 +/-3.8 years; 91% White), defined by a VAF threshold of 2% (standard CHIP) or 8% (large CHIP). 20.4% of women had CHIP using the VAF ≥2% definition while 6.0% had CHIP using the VAF ≥8% definition. The most common CHIP type for both definitions was *DNMT3A* followed by *TET2* (**Table 1**). For both the VAF ≥2% and VAF ≥8% CHIP definitions, participants with CHIP were older and tended to have less follow-up time. During follow-up, 1,776 women developed the composite MCI/dementia outcome with a mean (standard deviation [SD]) follow-up time of 12.0 (6.7) years, and 660 developed the more restrictive outcome of adjudicated dementia with a mean (SD) follow-up time of 11.6 (6.6) years. Notably the cumulative incidence proportion and cumulative incidence rates of adjudicated dementia were lower among large CHIP VAF ≥8% carriers than large CHIP non-carriers (**Table 1**).

**Table 1:** Demographic/Clinical characteristics and CHIP.

| Characteristic | 2% VAF |  |  | 8% VAF |  |  |
| --- | --- | --- | --- | --- | --- | --- |
|  | No CHIP | CHIP | p | No CHIP | CHIP | p |
| Sample Size, n (%) | 3927 (80%) | 1007 (20%) | -- | 4638 (94.0%) | 296 (6.0%) | -- |
| Sequencing batch*, n (%) |  |  |  |  |  |  |
| 1 | 2353 (59.9%) | 635 (63.1%) | .071 | 2796 (60.3%) | 192 (64.9%) | .125 |
| 2 | 1574 (40.1%) | 372 (36.9%) |  | 1842 (39.7%) | 104 (35.1%) |  |
| Hormone Treatment Arm*, n (%) |  |  |  |  |  |  |
| E-alone intervention | 725 (18.5%) | 212 (21.1%) | .029 | 876 (18.9%) | 61 (20.6%) | .266 |
| E-alone control | 761 (19.4%) | 200 (19.9%) |  | 900 (19.4%) | 61 (20.6%) |  |
| E+P intervention | 1215 (30.9%) | 266 (26.4%) |  | 1407 (30.3%) | 74 (25.0%) |  |
| E+P control | 1226 (31.2%) | 329 (32.7%) |  | 1455 (31.4%) | 100 (33.8%) |  |
| Age*, mean (SD) | 69.7 (3.7) | 70.5 (3.9) | <.001 | 69.9 (3.7) | 70.5 (4.0) | .006 |
| Age Bin*, n (%) |  |  |  |  |  |  |
| 60-64 | 98 (2.5%) | 17 (1.7%) | <.001 | 109 (2.4%) | 6 (2.0%) | .012 |
| 65-69 | 2001 (51.0%) | 434 (43.1%) |  | 2310 (49.8%) | 125 (42.2%) |  |
| 70-74 | 1317 (33.5%) | 377 (37.4%) |  | 1588 (34.2%) | 106 (35.8%) |  |
| 75-79 | 511 (13.0%) | 179 (17.8%) |  | 631 (13.6%) | 59 (19.9%) |  |
| Race and Ethnicity*, n (%) |  |  |  |  |  |  |
| White | 3562 (90.7%) | 923 (91.7%) | .365 | 4214 (90.9%) | 271 (91.6%) | .503 |
| Black/African American | 297 (7.6%) | 64 (6.4%) |  | 343 (7.4%) | 18 (6.1%) |  |
| Other | 68 (1.7%) | 20 (2.0%) |  | 81 (1.7%) | 7 (2.4%) |  |
| Education, n (%) |  |  |  |  |  |  |
| Up to 11 years | 264 (6.7%) | 73 (7.2%) | .084 | 309 (6.7%) | 28 (9.5%) | .038 |
| High school diploma/GED/vocational | 1331 (33.9%) | 341 (33.9%) |  | 1578 (34.0%) | 94 (31.8%) |  |
| Some college or associate degree | 1113 (28.3%) | 249 (24.7%) |  | 1295 (27.9%) | 67 (22.6%) |  |
| College graduate | 1219 (31.0%) | 344 (34.2%) |  | 1456 (31.4%) | 107 (36.1%) |  |
| US Region*, n (%) |  |  |  |  |  |  |
| Northeast | 1075 (27.4%) | 298 (29.6%) | .061 | 1285 (27.7%) | 88 (29.7%) | .072 |
| South | 869 (22.1%) | 201 (20.0%) |  | 1018 (21.9%) | 52 (17.6%) |  |
| Midwest | 995 (25.3%) | 229 (22.7%) |  | 1159 (25.0%) | 65 (22.0%) |  |
| West | 988 (25.2%) | 279<br>(27.7%) |  | 1176 (25.4%) | 91 (30.7%) |  |
| BMI, mean (SD) | 28.6 (5.6) | 28.2 (5.5) | .047 | 28.6 (5.6) | 28.3 (5.7) | .435 |
| BMI Missing, n (%) | 18 (0.5%) | 5 (0.5%) |  | 22 (0.5%) | 1 (0.3%) |  |
| High Chol. Requiring Pills, n (%) |  |  |  |  |  |  |
| No | 3158 (80.4%) | 829<br>(82.3%) | .392 | 3740 (80.6%) | 247<br>(83.4%) | .201 |
| Yes | 711 (18.1%) | 166<br>(16.5%) |  | 829 (17.9%) | 48 (16.2%) |  |
| Missing | 58 (1.5%) | 12 (1.2%) |  | 69 (1.5%) | 1 (0.3%) |  |
| Hypertension Requiring Pills, n (%) |  |  |  |  |  |  |
| No | 2398 (61.1%) | 637<br>(63.3%) | .388 | 2841 (61.3%) | 194<br>(65.5%) | .344 |
| Yes | 1482 (37.7%) | 357<br>(35.5%) |  | 1740 (37.5%) | 99 (33.4%) |  |
| Missing | 47 (1.2%) | 13 (1.3%) |  | 57 (1.2%) | 3 (1.0%) |  |
| Baseline Diabetes,n (%) |  |  |  |  |  |  |
| No | 3690 (94.0%) | 945<br>(93.8%) | .198 | 4357 (93.9%) | 278<br>(93.9%) | .604 |
| Yes | 232 (5.9%) | 58 (5.8%) |  | 273 (5.9%) | 17 (5.7%) |  |
| Missing | 5 (0.1%) | 4 (0.4%) |  | 8 (0.2%) | 1 (0.3%) |  |
| Baseline CVD, n (%) |  |  |  |  |  |  |
| None | 3332 (84.8%) | 841<br>(83.5%) | .595 | 3926 (84.6%) | 247<br>(83.4%) | .776 |
| Stroke or TIA | 143 (3.6%) | 39 (3.9%) |  | 172 (3.7%) | 10 (3.4%) |  |
| Other CVD*** | 373 (9.5%) | 101<br>(10.0%) |  | 443 (9.6%) | 31 (10.5%) |  |
| Missing | 79 (2.0%) | 26 (2.6%) |  | 97 (2.1%) | 8 (2.7%) |  |
| Alcohol Use, n (%) |  |  |  |  |  |  |
| <1 drink per month | 1750 (44.6%) | 442<br>(43.9%) | .830 | 2070 (44.6%) | 122<br>(41.2%) | .376 |
| >= 1 drink per month | 2167 (55.2%) | 562<br>(55.8%) |  | 2556 (55.1%) | 173<br>(58.4%) |  |
| Missing | 10 (0.3%) | 3 (0.3%) |  | 12 (0.3%) | 1 (0.3%) |  |
| Smoking Status, n (%) |  |  |  |  |  |  |
| Never | 2060 (52.5%) | 526<br>(52.2%) | .424 | 2445 (52.7%) | 141<br>(47.6%) | .077 |
| Former | 1555 (39.6%) | 408<br>(40.5%) |  | 1830 (39.5%) | 133<br>(44.9%) |  |
| Current | 262 (6.7%) | 56 (5.6%) |  | 303 (6.5%) | 15 (5.1%) |  |
| Missing | 50 (1.3%) | 17 (1.7%) |  | 60 (1.3%) | 7 (2.4%) |  |
| APOE2 Allele(s), n (%) |  |  |  |  |  |  |
| 0 | 3017 (76.8%) | 757<br>(75.2%) | .216 | 3544 (76.4%) | 230<br>(77.7%) | .203 |
| 1 | 539 (13.7%) | 138<br>(13.7%) |  | 642 (13.8%) | 35 (11.8%) |  |
| 2 | 18 (0.5%) | 9 (0.9%) |  | 23 (0.5%) | 4 (1.4%) |  |
| Missing | 353 (9.0%) | 103<br>(10.2%) |  | 429 (9.2%) | 27 (9.1%) |  |
| APOE4 Allele(s), n (%) |  |  |  |  |  |  |
| 0 | 2624 (66.8%) | 666 (66.1%) | .363 | 3099 (66.8%) | 191 (64.5%) | .046 |
| 1 | 885 (22.5%) | 216 (21.4%) |  | 1035 (22.3%) | 66 (22.3%) |  |
| 2 | 65 (1.7%) | 22 (2.2%) |  | 75 (1.6%) | 12 (4.1%) |  |
| Missing | 353 (9.0%) | 103 (10.2%) |  | 429 (9.2%) | 27 (9.1%) |  |
| APOE Risk, n (%) |  |  |  |  |  |  |
| low | 461 (11.7%) | 127 (12.6%) | .504 | 558 (12.0%) | 30 (10.1%) | .687 |
| neutral | 2163 (55.1%) | 539 (53.5%) |  | 2541 (54.8%) | 161 (54.4%) |  |
| high | 950 (24.2%) | 238 (23.6%) |  | 1110 (23.9%) | 78 (26.4%) |  |
| Missing | 353 (9.0%) | 103 (10.2%) |  | 429 (9.2%) | 27 (9.1%) |  |
| CHIP Type, n (%) |  |  |  |  |  |  |
| No CHIP | 3927 (100.0%) | 0 (0.0%) | -- | 4638 (100.0%) | 0 (0.0%) | -- |
| DNMT3A | - | 596 (59.2%) |  | - | 165 (55.7%) |  |
| TET2 | - | 167 (16.6%) |  | - | 70 (23.6%) |  |
| Other CHIP | - | 162 (16.1%) |  | - | 51 (17.2%) |  |
| Multi CHIP | - | 82 (8.1%) |  | - | 10 (3.4%) |  |
| MCI/PD Specific Demographics |  |  |  |  |  |  |
| MCI/PD, n (CIP) | 1407 (35.8%) | 359 (35.7%) | 0.941 | 1664 (35.9%) | 102 (34.5%) | 0.662 |
| Total Person Years | 47691.7 | 11710.9 | -- | 56004.9 | 3397.7 | -- |
| Cumulative Incidence Rate per 100 PY | 3 | 3.1 | 0.517 | 3 | 3 | 0.919 |
| Person Time, mean (SD) | 12.1 (6.8) | 11.6 (6.6) | < .001 | 12.1 (6.8) | 11.5 (6.4) | < .001 |
| PD Specific Demographics |  |  |  |  |  |  |
| PD, n (CIP) | 513 (13.1%) | 140 (13.9%) | 0.498 | 624 (13.5%) | 29 (9.8%) | 0.077 |
| Total Person Years | 46133.4 | 11212.1 | -- | 54190.4 | 3155.1 | -- |
| Cumulative Incidence Rate per 100 PY | 1.1 | 1.2 | 0.224 | 1.2 | 0.9 | 0.234 |
| Person Time, mean (SD) | 11.7 (6.7) | 11.1 (6.4) | < .001 | 11.7 (6.6) | 10.7 (6.1) | < .001 |

When CHIP was defined using VAF ≥2%, there was no significant difference in risk of cognitive impairment for either the broader definition of MCI/dementia or the narrower definition of adjudicated dementia (all *p-*values ≥0.1) (**Table 2**). By contrast, we observed a suggestive association of the presence of larger CHIP clone size (VAF ≥8%) with lower incidence of adjudicated dementia, whereas the HR using the broader definition of cognitive impairment was closer to 1.0 and not significant (**Table 2**). In the fully adjusted model (M3), the lower risk of developing adjudicated dementia associated with large CHIP (VAF ≥8%) was nominally significant (M3 HR = 0.62, 95%CI 0.41 - 0.94, p = 0.025). The protective association between large CHIP and probable dementia was weaker and not significant in the models that did not include APOE genotypes as covariates (M1, M2, **Table 2**), even when the M1 and M2 analyses were restricted to the same subset of participants included in M3 (**Table S1**).

**Table 2:** Association cognitive phenotypes and CHIP.

| **Mild Cognitive Impairment & Adjudicated Probable Dementia** |  |  |  |  |  |
| --- | --- | --- | --- | --- | --- |
|  | **HR** | **CI** | **N** | **CHIP** | **p** |
| **2% CHIP M1** | 0.96 | [0.86, 1.08] | 4934 | 1007 | .526 |
| **2% CHIP M2** | 0.95 | [0.84, 1.07] | 4655 | 943 | .398 |
| **2% CHIP M3** | 0.95 | [0.84, 1.08] | 4240 | 850 | .466 |
| **8% CHIP M1** | 0.99 | [0.81, 1.21] | 4934 | 296 | .896 |
| **8% CHIP M2** | 0.96 | [0.78, 1.18] | 4655 | 278 | .700 |
| **8% CHIP M3** | 0.89 | [0.72, 1.11] | 4240 | 254 | .293 |
| **Adjudicated Probable Dementia\*** |  |  |  |  |  |
|  | **HR** | **CI** | **N** | **CHIP** | **p** |
| **2% CHIP M1** | 1.04 | [0.86, 1.25] | 3821 | 223 | .702 |
| **2% CHIP M2** | 1.02 | [0.83, 1.24] | 3601 | 208 | .855 |
| **2% CHIP M3** | 1.03 | [0.84, 1.26] | 3236 | 188 | .787 |
| **8% CHIP M1** | 0.88 | [0.60, 1.27] | 3821 | 223 | .485 |
| **8% CHIP M2** | 0.79 | [0.53, 1.18] | 3601 | 208 | .251 |
| **8% CHIP M3** | 0.62 | [0.41, 0.94] | 3236 | 188 | **.025** |
| Survival analysis. The table contains adjusted hazard ratios (HR), confidence intervals (CI), sample size (N), number of CHIP positive (CHIP), and p-values (p). M1 covariates: age at blood draw, sequencing batch, hormone therapy, race/ethnicity, US region. M2 covariates: covariates in M1 + education level, alcohol use, smoking, BMI, cholesterol, hypertension, and CVD. M3 covariates: M2 + APOE2, APOE4.<br>*individuals with mild cognitive impairment were excluded from the analysis. |  |  |  |  |  |

Sensitivity analyses with death as a competing risk showed similar results to the survival model, confirming the association between CHIP and adjudicated probable dementia for larger CHIP clones VAF ≥8% (M3 HR = 0.61, p = 0.023; **Table S2**). Next, CHIP was modeled as a 3-category variable using VAF thresholds of <2%, 2% to 8%, and ≥8% (No CHIP, low and large CHIP clones). In fully adjusted M3, large CHIP clones (VAF ≥8%) had a nominally significant association with adjudicated probable dementia, (HR = 0.64, p = 0.036; **Table S3**), but there was no evidence of a protective association for the smaller clone category.

To explore differences in the protective association of large CHIP clones on probable dementia by APOE genotype, the large CHIP VAF ≥8% analysis was repeated stratifying participants into three APOE risk categories [neutral (*APOE* ε3ε3), low-risk (*APOE* ε2ε2 and ε2ε3) and high-risk (any *APOE* ε4 allele)]. The apparent protective association of CHIP was strongest in the high-risk group (HR = 0.58, 95% CI 0.31-1.10, p = 0.094; **Table 3**) and progressively closer to the null in the neutral and low-risk APOE groups.

**Table 3:**
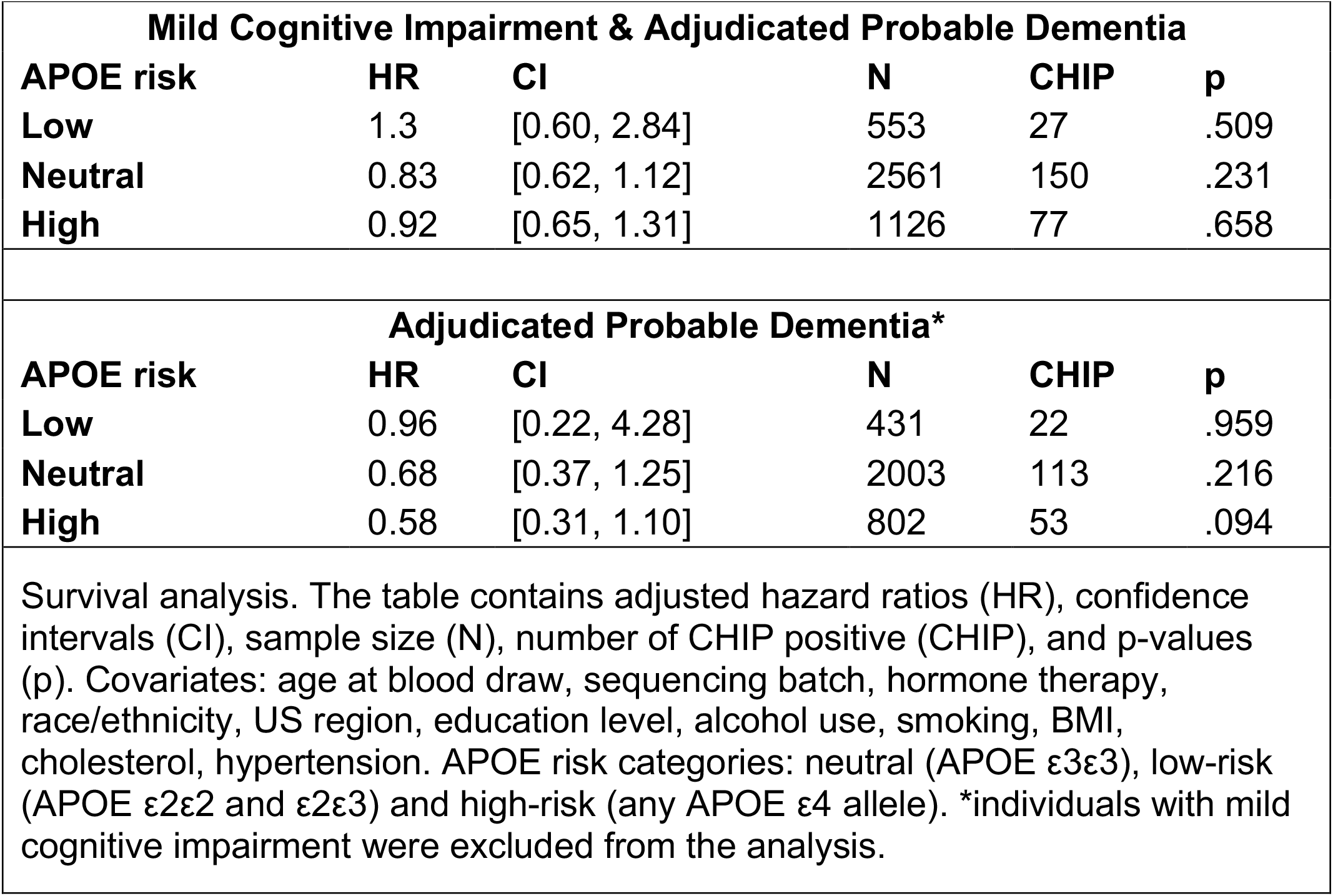
Association cognitive phenotypes and CHIP 8% with stratified APOE risk.

| Mild Cognitive Impairment & Adjudicated Probable Dementia |  |  |  |  |  |
| --- | --- | --- | --- | --- | --- |
| APOE risk | HR | CI | N | CHIP | p |
| Low | 1.3 | [0.60, 2.84] | 553 | 27 | .509 |
| Neutral | 0.83 | [0.62, 1.12] | 2561 | 150 | .231 |
| High | 0.92 | [0.65, 1.31] | 1126 | 77 | .658 |
| Adjudicated Probable Dementia* |  |  |  |  |  |
| APOE risk | HR | CI | N | CHIP | p |
| Low | 0.96 | [0.22, 4.28] | 431 | 22 | .959 |
| Neutral | 0.68 | [0.37, 1.25] | 2003 | 113 | .216 |
| High | 0.58 | [0.31, 1.10] | 802 | 53 | .094 |
| Survival analysis. The table contains adjusted hazard ratios (HR), confidence intervals (CI), sample size (N), number of CHIP positive (CHIP), and p-values (p). Covariates: age at blood draw, sequencing batch, hormone therapy, race/ethnicity, US region, education level, alcohol use, smoking, BMI, cholesterol, hypertension. APOE risk categories: neutral (APOE $\epsilon 3\epsilon 3$), low-risk (APOE $\epsilon 2\epsilon 2$ and $\epsilon 2\epsilon 3$) and high-risk (any APOE $\epsilon 4$ allele). *individuals with mild cognitive impairment were excluded from the analysis. | | | | | |

Next, we explored the association between baseline CHIP and incident cognitive impairment according to CHIP driver gene subtype. In the fully adjusted model (**Table 4**), the protective association of large CHIP (VAF ≥8%) on risk of incident adjudicated dementia was apparent among carriers of *DNMT3A* mutations (M3 HR = 0.48, 95% CI 0.27-0.86, *p* = 0.014) but not among *TET2* mutation carriers (M3 HR = 1.11, 95% CI 0.55-2.26, *p* = 0.77). However, these results are exploratory and not corrected for multiple comparisons. Hazards for developing the composite MCI/dementia outcome using the 2% VAF CHIP definition trended in the same direction, but did not reach significance (**Table S4**).

**Table 4:** Association of adjudicated probable dementia and CHIP mutations.

|  | **HR** | **CI** | **N** | **CHIP** | **p** |
| --- | --- | --- | --- | --- | --- |
| **DNMT3A** | 0.48 | [0.27, 0.86] | 3236 | 110 | **.014** |
| **TET2** | 1.11 | [0.55, 2.26] | 3236 | 44 | .768 |
| **Other** | 0.65 | [0.24, 1.76] | 3236 | 29 | .399 |
| **Multiple** | 0.00 | [0.00, NA] | 3236 | 5 | .988 |

Survival analysis. The table contains adjusted hazard ratios (HR), confidence intervals (CI), sample size (N), number of CHIP positive (CHIP), and p-values (p). Data shown for M3 covariates: age at blood draw, sequencing batch, hormone therapy, race/ethnicity, US region, education level, alcohol use, smoking, BMI, cholesterol, hypertension, CVD, APOE2, APOE4. Individuals with mild cognitive impairment were excluded from the analysis.

## Discussion

Utilizing data from the prospective, community-based WHIMS, we found that larger CHIP clones (≥8% VAF) were nominally associated with lower risk of incident adjudicated dementia in postmenopausal women when adjusting for traditional risk factors including APOE genotype. The direction of effect is consistent with two recent observational studies indicating a protective association of CHIP on AD[13] or impairment of attention and executive cognitive function among CKD patients[9]. Given the well-established relationship between CHIP and *increased* risk of other chronic aging-related and inflammatory diseases (atherosclerotic CVD, stroke, and mortality), the protective association of CHIP on neurodegenerative outcomes initially appears counter-intuitive. However, largely consistent findings of a protective association have now been observed across at least three human cohort studies and are further supported by Mendelian randomization and functional studies of *postmortem* brain tissue demonstrating the role of mutant, marrow-derived cells in reducing neuritic plaques and neurofibrillary tangles[13]. Other age-related conditions characterized by non-malignant clonal expansion of hematopoietic cells driven by myeloid malignancy-associated somatic driver mutations (e.g., myelodysplastic syndrome) have similarly been associated with reduced risk of neurodegenerative diseases including dementia[14].

In WHIMS, the protective association of CHIP on adjudicated dementia was observed for large CHIP clones (VAF ≥8%), but there was little evidence for association when CHIP was defined using the traditional VAF threshold of ≥2%. Moreover, the association of large CHIP clones with the combined outcome that includes both dementia and milder cognitive deficits was weaker and not statistically significant. These findings are analogous to those reported in the Alzheimer’s Disease Sequencing Project (ADSP)[13], in which CHIP with VAF >8% was associated with a stronger protective association of AD dementia (OR = 0.66, *P* = 5.5 × 10^−4^) than using a CHIP VAF cutoff of 2% (OR = 0.79, *P* = 0.024), whereas a VAF ≤8% had no association with dementia in ADSP (OR = 1.25, *P* = 0.23). CHIP clone size has previously been shown to be an important predictor of risk for other aging-related disorders including hematologic malignancy and cardiovascular outcomes[3,4,15]. In contrast, the lower risk of cognitive impairment phenotypes among CKD patients was confined to those with small CHIP clone size, whereas no association was observed for large CHIP clone size[9]. The reasons for the lack of dose-response relationship in the latter CKD study are unclear. It is possible that differences in study design, participant characteristics (including presence of underlying CKD), sample sizes, VAF distribution, or cognitive outcome definitions may contribute to some of the differences in findings observed across the three studies.

When CHIP was analyzed according to major driver gene subtypes in WHIMS, associations with cognitive impairment were stronger across *DNMT3A* than *TET2* CHIP, a pattern that was also observed by Xiao et al in CKD patients[9]. In contrast, consistent protective associations of CHIP driver genes, including *DNMT3A, TET2, ASXL1*, and *SF3B1*, were observed on AD[13].

When stratified by *APOE* risk alleles, we observed a trend toward more protective associations among those without any e2 alleles, which is also consistent with previous observations[9,13]. Additional studies with larger sample sizes and sensitive CHIP detection assays are needed to clarify the role of both CHIP clone size, individual CHIP driver gene subtype, and APOE genotype on risk of cognitive impairment.

A strength of the current study is the length of follow-up in a community-based sample of older women who are more likely than men to experience adverse cognitive outcomes later in life. However, one limitation of this study is the lack of generalization of findings to men. Mosaic loss of chromosome Y is another type of clonal hematopoiesis common in older men, and has been associated with increased AD risk[16]. Larger studies including both men and women are needed to elucidate the combined effects of CHIP and other somatic mutations in hematopoietic cells on the risk of adverse cognitive outcomes. A second limitation of this study is the inability to distinguish between different etiologies of cognitive impairment, such as vascular dementia, AD, and other neurodegenerative conditions. In the UK Biobank, CHIP was associated with increased risk of vascular, but not with primary, neurodegenerative diseases[17]. In another study, CHIP with *TET2* mutations was associated with increased risk of Parkinson’s disease[18]. Therefore, the protective association of CHIP on cognitive impairment may be specific to certain neurodegenerative conditions such as AD. Additional clinical and population studies that include brain imaging and AD biomarkers are needed to better understand the specificity and mechanisms underlying these relationships.

## Data Availability

Data needs to be requested through the Womens Health Initiative Repository. Sequencing data used in this study is deposited in dbGaP (phs000200.v12.p3).

## Acknowledgments/Funding

This work was supported by NIH R01 grants HL148565 and HL148565-02S1 (A.P.R., E.A.W). The WHI program is funded by the National Heart, Lung, and Blood Institute, National Institutes of Health, U.S. Department of Health and Human Services through contracts 75N92021D00001, 75N92021D00002, 75N92021D00003, 75N92021D00004, 75N92021D00005.

