## Supplementary Text and Tables for "Clonal Hematopoiesis of Indeterminate Potential and the Risk of Cognitive Impairment in the Women’s Health Initiative Memory Study"

**Supplementary Information**

*Cognitive Assessment and Outcome Definitions*

The original WHIMS protocol included annual administration of the Modified Mini-Mental State Exam (3MS)[1]. Women scoring below pre-determined cut points (based on age and education) were referred for a clinical evaluation by a board-certified physician and neuropsychological testing, including portions of the CERAD battery, the Mini-Mental State Examination (MMSE), the Trail Making Test Parts A and B, a structured psychiatric interview (PRIME-MD), and the Geriatric Depression Scale short form (GDS). A knowledgeable informant completed the Acquired Cognitive and Behavior Changes (ACBD) form. A central panel of dementia experts then adjudicated cases, classifying participants as cognitively normal, MCI, or probable dementia, according to standardized criteria (DSM-IV). In WHIMS-ECHO, a validated telephone cognitive battery was administered annually to all participants[2]. The battery comprised a modified version of the Telephone Interview for Cognitive Status (TICS-m), the Oral Trail Making Tests Part A and B, the East Boston Memory Test, Digit Span, and verbal fluency/animals. When participants scored below 31 points on the TICS-m, the Dementia Questionnaire was administered to an informant to evaluate functional status. Cognitive status was then adjudicated as MCI, probable dementia, or no impairment, as described above. In addition to cognitive function testing performed in WHIMS/WHIMS-ECHO participants, another source of WHI data on dementia was obtained by self-report ("Has a doctor told you for the first time that you have moderate or severe memory problems, dementia, or Alzheimer's?"). The self-reported dementia question was administered annually starting in 2005 to all surviving WHI participants as part of outcomes adjudication for the WHI extension studies. Validation of WHI self-reported dementia/Alzheimer's data using either adjudicated WHIMS probable dementia/MCI or Medicare claims data showed very high specificity (97% and 95%, respectively) and PPV (72% and 75%, respectively).

*CHIP assay and Definition of CHIP Status at Baseline*

For a subset of 4,949 WHIMS women who provided blood samples at the WHI baseline exam, genomic DNA extracted from stored buffy coats were shipped to the University of Michigan for evaluation of CHIP using targeted sequencing. A smMIPS capture panel was designed to tile coding exons (+/− 5 bp) of the 11 most common CHIP genes and recurrent mutational hotspots in four others, as previously described[3,4]. Sequencing reads were aligned to the human reference genome (build 37), and a custom sequencing pipeline (https://github.com/kitzmanlab/mimips) was used for post-alignment processing. Somatic SNPs and indels were called using LoFreq 2.1.3.1,[5] requiring minimum coverage 40, with ≥5 reads supporting the alternate allele and a variant allele frequency (VAF) ≥0.1%. Variants present in ≥5% of samples at a VAF of 1–10% were discarded as likely recurrent artifacts. Variants were annotated using ANNOVAR software[6] and processed using an existing filtering pipeline based upon gene name, variant functional class, and population allele frequency. The workflow is available at https://app.terra.bio/#workspaces/terra-outreach/CHIP-Detection-Mutect2/notebooks. We manually reviewed alignments for selected CHIP variant calls using Integrative Genomics Viewer (IGV).

**Supplementary Table 1:** Association cognitive phenotypes and CHIP for subset of women with complete data across all covariates in model 3

| | HR | CI | N*** | CHIP | p |
|---|---|---|---|---|---|
| **Mild Cognitive Impairment & Adjudicated Probable Dementia** | | | | | |
| **2% CHIP M1** | 0.95 | [0.84, 1.08] | 4240 | 850 | .463 |
| **2% CHIP M2** | 0.94 | [0.83, 1.07] | 4240 | 850 | .335 |
| **2% CHIP M3** | 0.95 | [0.84, 1.08] | 4240 | 850 | .466 |
| **8% CHIP M1** | 0.95 | [0.77, 1.18] | 4240 | 254 | .647 |
| **8% CHIP M2** | 0.94 | [0.76, 1.16] | 4240 | 254 | .547 |
| **8% CHIP M3** | 0.89 | [0.72, 1.11] | 4240 | 254 | .293 |

| | HR | CI | N*** | CHIP | p |
|---|---|---|---|---|---|
| **Adjudicated Probable Dementia*** | | | | | |
| **2% CHIP M1** | 0.99 | [0.81, 1.22] | 3236 | 188 | .959 |
| **2% CHIP M2** | 0.99 | [0.81, 1.22] | 3236 | 188 | .959 |
| **2% CHIP M3** | 1.03 | [0.84, 1.26] | 3236 | 188 | .787 |
| **8% CHIP M1** | 0.77 | [0.51, 1.16] | 3236 | 188 | .207 |
| **8% CHIP M2** | 0.74 | [0.49, 1.11] | 3236 | 188 | .148 |
| **8% CHIP M3** | 0.62 | [0.41, 0.94] | 3236 | 188 | **.025** |

Survival analysis. The table contains adjusted hazard ratios (HR), confidence intervals (CI), sample size (N), number of CHIP positive (CHIP), and p-values (p). N*** The same subset of participants were used across models, those with complete data for all covariates in M3. M1 covariates: age at blood draw, sequencing batch, hormone therapy, race/ethnicity, US region. M2 covariates: covariates in M1 + education level, alcohol use, smoking, BMI, cholesterol, hypertension, and CVD. M3 covariates: M2 + APOE2, APOE4. *individuals with mild cognitive impairment were excluded from the analysis.

**Supplementary Table 2:** Association cognitive phenotypes and CHIP with death as a competing risk

| | HR | CI | N | CHIP | p |
|---|---|---|---|---|---|
| **Mild Cognitive Impairment & Adjudicated Probable Dementia** | | | | | |
| | **HR** | **CI** | **N** | **CHIP** | **p** |
| **2% CHIP M1** | 0.98 | [0.88, 1.10] | 4925 | 1004 | .755 |
| **2% CHIP M2** | 0.98 | [0.87, 1.11] | 4646 | 940 | .792 |
| **2% CHIP M3** | 0.98 | [0.87, 1.11] | 4231 | 847 | .766 |
| **8% CHIP M1** | 0.96 | [0.81, 1.15] | 4925 | 295 | .688 |
| **8% CHIP M2** | 0.92 | [0.75, 1.13] | 4646 | 277 | .430 |
| **8% CHIP M3** | 0.87 | [0.70, 1.08] | 4231 | 253 | .204 |
| **Adjudicated Probable Dementia*** | | | | | |
| | **HR** | **CI** | **N** | **CHIP** | **p** |
| **2% CHIP M1** | 1.04 | [0.86, 1.26] | 3812 | 785 | .658 |
| **2% CHIP M2** | 1.04 | [0.85, 1.27] | 3592 | 732 | .698 |
| **2% CHIP M3** | 1.03 | [0.84, 1.28] | 3227 | 650 | .750 |
| **8% CHIP M1** | 0.81 | [0.56, 1.16] | 3812 | 222 | .252 |
| **8% CHIP M2** | 0.75 | [0.51, 1.10] | 3592 | 207 | .145 |
| **8% CHIP M3** | 0.61 | [0.40, 0.93] | 3227 | 187 | **.023** |

Competing risk analysis. The table contains adjusted hazard ratios (HR), confidence intervals (CI), sample size (N), number of CHIP positive (CHIP), and p-values (p). M1 covariates: age at blood draw, sequencing batch, hormone therapy, race/ethnicity, US region. M2 covariates: covariates in M1 + education level, alcohol use, smoking, BMI, cholesterol, hypertension, and CVD. M3 covariates: M2 + APOE2, APOE4. *individuals with mild cognitive impairment were excluded from the analysis.

**Supplementary Table 3:** Association cognitive phenotypes and CHIP modeling CHIP as a categorical variable: none, low, high

**Mild Cognitive Impairment & Adjudicated Probable Dementia**

| | CHIP category | HR | CI | N*** | CHIP | p |
|---|---|---|---|---|---|---|
| **Model 1** | 2-8% | 0.96 | [0.84, 1.09] | 4934 | 711 | .515 |
| **Model 1** | 8+% | 0.98 | [0.80, 1.20] | 4934 | 296 | .840 |
| **Model 2** | 2-8% | 0.95 | [0.82, 1.09] | 4655 | 665 | .453 |
| **Model 2** | 8+% | 0.95 | [0.77, 1.17] | 4655 | 278 | .641 |
| **Model 3** | 2-8% | 0.98 | [0.85, 1.14] | 4240 | 596 | .819 |
| **Model 3** | 8+% | 0.89 | [0.72, 1.10] | 4240 | 254 | .285 |

**Adjudicated Probable Dementia***

| | CHIP category | HR | CI | N*** | CHIP | p |
|---|---|---|---|---|---|---|
| **Model 1** | 2-8% | 1.09 | [0.88, 1.34] | 3821 | 711 | .436 |
| **Model 1** | 8+% | 0.89 | [0.61, 1.29] | 3821 | 296 | .536 |
| **Model 2** | 2-8% | 1.10 | [0.88, 1.36] | 3601 | 665 | .416 |
| **Model 2** | 8+% | 0.81 | [0.54, 1.20] | 3601 | 278 | .287 |
| **Model 3** | 2-8% | 1.21 | [0.96, 1.51] | 3236 | 596 | .101 |
| **Model 3** | 8+% | 0.64 | [0.42, 0.97] | 3236 | 254 | **.036** |

Survival analysis. The table contains adjusted hazard ratios (HR), confidence intervals (CI), sample size (N), number of CHIP positive (CHIP), and p-values (p). Chip was modeled as a categorical variable based on the variant allele frequency (VAF) of CHIP mutations in a sample. No chip was defined as <2% VAF, low chip between 2%-8% VAF and high chip as >8% VAF. For individuals with mutliple mutations the mutation with the highest VAF as used for clasification. The no chip group was used as the baseline. M1 covariates: age at blood draw, sequencing batch, hormone therapy, race/ethnicity, US region. M2 covariates: covariates in M1 + education level, alcohol use, smoking, BMI, cholesterol, hypertension, and CVD. M3 covariates: M2 + APOE2, APOE4. *individuals with mild cognitive impairment were excluded from the analysis.

| **Supplementary Table 4:** Association cognitive phenotypes and CHIP mutations | | | | | | |
|---|---|---|---|---|---|---|
| | | | **Mild Cognitive Impairment & Adjudicated Probable Dementia** | | | |
| | **CHIP Type** | **HR** | **CI** | **N** | **CHIP** | **p** |
| **2% CHIP M1** | DNMT3A | 0.95 | [0.82, 1.10] | 4934 | 596 | .483 |
| **2% CHIP M1** | TET2 | 1.02 | [0.79, 1.31] | 4934 | 167 | .889 |
| **2% CHIP M1** | Other | 1.07 | [0.82, 1.39] | 4934 | 162 | .608 |
| **2% CHIP M1** | Multiple | 0.78 | [0.54, 1.14] | 4934 | 82 | .199 |
| **2% CHIP M2** | DNMT3A | 0.94 | [0.81, 1.10] | 4655 | 561 | .434 |
| **2% CHIP M2** | TET2 | 1.02 | [0.79, 1.33] | 4655 | 154 | .874 |
| **2% CHIP M2** | Other | 1.07 | [0.81, 1.41] | 4655 | 148 | .649 |
| **2% CHIP M2** | Multiple | 0.72 | [0.49, 1.06] | 4655 | 80 | .096 |
| **2% CHIP M3** | DNMT3A | 0.96 | [0.82, 1.12] | 4240 | 502 | .590 |
| **2% CHIP M3** | TET2 | 1.01 | [0.77, 1.32] | 4240 | 142 | .951 |
| **2% CHIP M3** | Other | 0.97 | [0.72, 1.31] | 4240 | 133 | .850 |
| **2% CHIP M3** | Multiple | 0.81 | [0.55, 1.20] | 4240 | 73 | .298 |
| **8% CHIP M1** | DNMT3A | 0.89 | [0.67, 1.17] | 4934 | 165 | .405 |
| **8% CHIP M1** | TET2 | 1.06 | [0.73, 1.56] | 4934 | 70 | .743 |
| **8% CHIP M1** | Other | 1.21 | [0.79, 1.86] | 4934 | 51 | .388 |
| **8% CHIP M1** | Multiple | 0.89 | [0.22, 3.58] | 4934 | 10 | .874 |
| **8% CHIP M2** | DNMT3A | 0.87 | [0.65, 1.17] | 4655 | 157 | .353 |
| **8% CHIP M2** | TET2 | 1.05 | [0.72, 1.53] | 4655 | 65 | .809 |
| **8% CHIP M2** | Other | 1.11 | [0.71, 1.76] | 4655 | 46 | .640 |
| **8% CHIP M2** | Multiple | 0.90 | [0.22, 3.61] | 4655 | 10 | .881 |
| **8% CHIP M3** | DNMT3A | 0.81 | [0.60, 1.09] | 4240 | 144 | .157 |
| **8% CHIP M3** | TET2 | 1.03 | [0.70, 1.51] | 4240 | 63 | .897 |
| **8% CHIP M3** | Other | 0.94 | [0.56, 1.57] | 4240 | 40 | .811 |
| **8% CHIP M3** | Multiple | 1.15 | [0.28, 4.60] | 4240 | 7 | .849 |
| | | | **Adjudicated Probable Dementia*** | | | |
| | **CHIP Type** | **HR** | **CI** | **N** | **CHIP** | **p** |
| **2% CHIP M1** | DNMT3A | 1.00 | [0.78, 1.28] | 3821 | 465 | .995 |
| **2% CHIP M1** | TET2 | 1.20 | [0.81, 1.76] | 3821 | 130 | .367 |
| **2% CHIP M1** | Other | 1.14 | [0.76, 1.70] | 3821 | 128 | .534 |
| **2% CHIP M1** | Multiple | 0.81 | [0.44, 1.48] | 3821 | 65 | .500 |
| **2% CHIP M2** | DNMT3A | 1.02 | [0.79, 1.31] | 3601 | 438 | .895 |
| **2% CHIP M2** | TET2 | 1.11 | [0.73, 1.70] | 3601 | 117 | .628 |
| **2% CHIP M2** | Other | 1.15 | [0.74, 1.78] | 3601 | 117 | .549 |
| **2% CHIP M2** | Multiple | 0.72 | [0.38, 1.36] | 3601 | 63 | .318 |
| **2% CHIP M3** | DNMT3A | 1.10 | [0.85, 1.42] | 3236 | 386 | .477 |
| **2% CHIP M3** | TET2 | 1.08 | [0.70, 1.68] | 3236 | 107 | .730 |
| **2% CHIP M3** | Other | 0.90 | [0.55, 1.47] | 3236 | 104 | .663 |
| **2% CHIP M3** | Multiple | 0.80 | [0.42, 1.51] | 3236 | 56 | .487 |
| **8% CHIP M1** | DNMT3A | 0.66 | [0.38, 1.15] | 3821 | 127 | .144 |
| **8% CHIP M1** | TET2 | 1.19 | [0.59, 2.40] | 3821 | 50 | .625 |
| **8% CHIP M1** | Other | 1.36 | [0.67, 2.75] | 3821 | 38 | .388 |
| **8% CHIP M1** | Multiple | 0.00 | [0.00, NA] | 3821 | 8 | .985 |
| **8% CHIP M2** | DNMT3A | 0.63 | [0.35, 1.11] | 3601 | 122 | .112 |
| **8% CHIP M2** | TET2 | 1.14 | [0.56, 2.30] | 3601 | 45 | .721 |
| **8% CHIP M2** | Other | 1.06 | [0.47, 2.40] | 3601 | 33 | .888 |
| **8% CHIP M2** | Multiple | 0.00 | [0.00, NA] | 3601 | 8 | .985 |
| **8% CHIP M3** | DNMT3A | 0.48 | [0.27, 0.86] | 3236 | 110 | **.014** |
| **8% CHIP M3** | TET2 | 1.11 | [0.55, 2.26] | 3236 | 44 | .768 |

| | | | | | | |
|---|---|---|---|---|---|---|
| **8% CHIP M3** | Other | 0.65 | [0.24, 1.76] | 3236 | 29 | .399 |
| **8% CHIP M3** | Multiple | 0.00 | [0.00, NA] | 3236 | 5 | .988 |

Survival analysis. The table contains adjusted hazard ratios (HR), confidence intervals (CI), sample size (N), number of CHIP positive (CHIP), and p-values (p). M1 covariates: age at blood draw, sequencing batch, hormone therapy, race/ethnicity, US region. M2 covariates: covariates in M1 + education level, alcohol use, smoking, BMI, cholesterol, hypertension, and CVD. M3 covariates: M2 + APOE2, APOE4. *individuals with mild cognitive impairment were excluded from the analysis.
